# Performance of five risk stratification tools for paediatric pneumonia against WHO scores using data from the PediCAP trial in sub-Saharan Africa

**DOI:** 10.64898/2026.06.02.26353886

**Authors:** Damalie Nalwanga, Michelle Clements, Victor Musiime, Veronica Mulenga, Hilda A Mujuru, Muhammad Sidat, W. Chris Buck, Shabir A. Madhi, Julia A Bielicki, David P. Moore, A Sarah Walker, Mike Sharland, the PediCAP trial team

## Abstract

**Objectives:** Risk stratification tools for childhood pneumonia may improve identification of children at highest risk of death in low-resource settings. However, added value over the WHO Integrated Management of Childhood Illness (IMCI) criteria and danger signs remains uncertain. We compared the performance of published paediatric pneumonia risk scores with WHO-IMCI criteria and WHO danger signs.

**Design:** Secondary analysis of a multi-country randomised controlled trial

**Setting:** Thirteen hospitals in Mozambique, South Africa, Uganda, Zambia, and Zimbabwe.

**Participants:** Children aged 2-59 months without HIV hospitalised with WHO-defined severe community-acquired pneumonia in the PediCAP trial. Children requiring intensive care or invasive ventilation were excluded.

**Primary and secondary outcome measures:** The primary outcome was in-hospital mortality. The exploratory outcome was readmission or death by day 28. Discrimination was assessed using area under the receiver operating characteristic curve (ROC-AUC).

**Results:** Among 1052 children, 18 (1.7%) died in hospital and 22 (2.1%) died in hospital or within 7 days of discharge. Among 1010 children with day-28 follow-up, 63 (6.2%) died or were readmitted by day 28. ROC-AUC point estimates for in-hospital mortality ranged from approximately 0.73 to 0.87. WHO danger signs had lower point estimate than the other scores, although confidence intervals overlapped substantially and no score demonstrated clear superiority. In contrast, all approaches showed limited discrimination for the exploratory outcome of 28-day readmission or death (ROC-AUC approximately 0.54-0.57).

**Conclusions:** In this multi-country cohort of children hospitalised with severe pneumonia, no score demonstrated clear superiority for mortality prediction, although point estimates of AUC-ROC scores ranged from 0.75 (Danger signs) to 0.87 (RISC-MGMU). Published risk scores and WHO-IMCI criteria showed broadly similar performance. These findings suggest that opportunities to improve risk stratification in low-resource settings may lie not only in developing new tools, but also in optimising the delivery and use of existing WHO IMCI-based assessment.

**Trial Registration:** ISRCTN63115131

**What is already known on this topic:** Several clinical risk scores have been developed to predict mortality in childhood pneumonia, with some validation studies in low-resource settings, although direct comparisons across multiple risk stratification tools within the same population are limited.

WHO IMCI criteria are widely used in many low-resource settings, but performance relative to published risk stratification tools remains uncertain.

**What this study adds:** In a multi-country African cohort of children hospitalised with severe pneumonia, published paediatric pneumonia risk scores and WHO IMCI criteria showed broadly similar point estimates for mortality prediction, with no score demonstrating clear superiority.

WHO danger signs generally had lower point estimates than the published risk scores and WHO IMCI criteria, although confidence intervals overlapped substantially.

**How this study might affect research, practice, or policy:** These findings suggest that opportunities to improve risk stratification in low-resource settings may lie not only in developing new tools, but also in optimising the delivery and use of existing WHO IMCI-based assessment.

## Introduction

Community-acquired pneumonia remains one of the most significant causes of morbidity and mortality among children under-5 years, resulting in over 725,000 deaths annually^1^. Most paediatric pneumonia deaths occur shortly after hospital admission^2 3^, although some deaths may occur following hospital discharge^4^. Identifying children who are at high risk of mortality from pneumonia is imperative, as it supports health workers to prioritise and monitor high-risk patients more closely. To this end, a number of risk scores have been developed for paediatric pneumonia^5^.

Scores for risk stratification in paediatric pneumonia have been developed using cohort and clinical trial data, and include information on medical history, clinical examination findings, and laboratory investigations^6-10^. While several risk scores have been developed and tested using data from the populations in which they were developed^11^, validation across populations outside the original sample is essential to ensure robustness and establish their more generalisable utility, yet it is more rarely performed. Furthermore, comparing the utility of these scores, particularly using more routinely available clinical data, such as history and clinical examination alone, is particularly beneficial to health workers in low- and middle-income countries (LMICs) where laboratory investigations may not be readily accessible.

Further, the World Health Organisation (WHO) has proposed severity indicator criteria for pneumonia and general danger signs applicable to all children being admitted to hospital^12^: how condition-specific scores relate to general risk scores has rarely been considered, despite the more widespread applicability of the latter.

This study, therefore, aimed to compare, using prospectively recorded data derived from a multi-country clinical trial, the performance and utility of published pneumonia risk-stratification tools with Integrated Management of Childhood Illnesses (IMCI) pneumonia signs and the WHO danger signs among children with severe pneumonia, who are at the highest risk of death.

## Methods

We used data from children 2-<60 months hospitalised for severe community acquired pneumonia, defined according to the WHO classification^13^ and judged to require at least 24 hours of intravenous antibiotics, who were recruited in the PediCAP randomised controlled trial conducted in 13 hospitals in Mozambique, South Africa, Uganda, Zambia, and Zimbabwe^14^ from December 2020 to August 2023. Children who had already received intravenous antibiotics for more than 24 hours, or were receiving outpatient oral antibiotics as prophylaxis, such as children living with HIV (CLHIV) receiving cotrimoxazole prophylaxis, were excluded from the trial, as were children with known or anticipated need for invasive ventilation or admission to intensive care. Other exclusion criteria included point-of-care semi-quantitative C-reactive protein (CRP) test < 10mg/l at screening (very unlikely to represent severe pneumonia requiring antibiotics), possible nosocomial pneumonia (onset > 48 hours after admission or admitted to hospital overnight within the last 28 days) and diagnosis likely not community acquired pneumonia (asthma or viral bronchiolitis in a child under 1 year of age). The PediCAP trial evaluated the effectiveness and safety of an oral step-down strategy to dispersible amoxicillin and co-amoxiclav with varying total durations of antibiotic treatment compared with 5 days of intravenous treatment for severe community-acquired pneumonia.

We evaluated risk stratification tools whose models were published (reviewed by Rees et al. 2025^5^ plus one other^15^), and whose parameters were available in PediCAP. Given the available data, risk stratification tools developed for outpatient populations, using laboratory data, developed for CLHIV, or that included parameters not assessed in PediCAP were not evaluated (**Table S1**). Five scores were therefore compared using data available at admission: PREPARE^6^, RISC-KGMU^7^, PERCH^8^, RISC-Malawi^9^, RISC^10^ (summarised in **Table S2** and presented in order of development from most recent to earliest). Additionally, we included the WHO danger signs and the combined WHO danger signs plus signs of severe pneumonia from the WHO IMCI^12^ as simple clinical comparators, given their widespread use in routine care, but lacking formal derivation as risk prediction tools. WHO danger signs and WHO IMCI severe pneumonia criteria were not treated as binary positive/negative classifications. Instead, one point was assigned for each danger sign or severity sign present and the total count was used as a score. The RISC-KGMU score was implemented as the published RISC score with weight-for-height z-score replacing weight-for-age z-score, consistent with the KGMU modification. PERCH, PREPARE, and RISC-Malawi scores were analysed including and excluding the score component of ‘unconscious’, as this was captured in PediCAP as ‘lethargy or unconsciousness’. All Z-scores were calculated using WHO norms^16^. All risk score components were assessed as soon as possible after randomisation and recorded according to the worst status observed since admission, ensuring that improvement following intravenous antibiotic treatment did not materially influence measurement.

The primary outcome was in-hospital mortality as this was the derivation outcome for four of the five evaluated scores. Mortality occurring during hospitalisation or within seven days of discharge was evaluated as a secondary outcome, as this was the derivation outcome for the PERCH score. Death or readmission by day 28 was included as an exploratory outcome as it was the primary outcome of the PediCAP trial and post-discharge mortality and readmission represent a substantial and under-recognised infection outcome in resource-constrained settings, including for pneumonia. Additionally, readmission contributes to the pneumonia burden in such settings and is highly relevant to patients, families, and health systems^17^. Readmission or death to day 28 was considered exploratory as it was not a derivation outcome for any of the evaluated risk scores.

### Statistical analysis

Analysis used only complete cases for all predictors and each outcome for comparability and because models did not specify a consistent approach to handling missing predictors. All scores were constructed so that higher scores represent higher risk. Univariate case-fatality ratios were calculated for each score component and outcome, with 95% confidence intervals calculated using the Wilson score method. The area under the receiver operating characteristic curve (ROC-AUC) was used to assess test performance, with associated 95% confidence intervals calculated using cluster bootstrap accounting for centre-level clustering (centres with fewer than five children grouped to the geographically nearest centre as per the primary analysis). Precision-recall area under the curve (PR-AUC) was also calculated given the low frequency of events. Performance statistics, sensitivity, specificity, and positive predictive value, were calculated with confidence intervals calculated by the exact binomial method. Analyses were performed in R version 4.5.1^18^. Code is available in **Code S1**)

### Patient and public involvement

Patients and the public were not involved in the design and conduct of this study.

### Ethics statement

Ethical approval was not required for this secondary analysis. For the main PediCAP trial, local ethical approval was sought and obtained in each country. We also obtained hospital or Provincial Health Authority approvals, in addition to regulatory approvals for the clinical trial (Mozambique – Comite Nacional de Bioethica Para a Saude (IRB00002657) REF 323/CNBS/22, South Africa - University of the Witwatersrand Human Research Ethics Committee (Medical) REF 190913B, Uganda – Makarere University Research Ethics Committee REF 2019-162, Zambia – University of Zambia Biomedical Research Ethics Committee REF 328-2019, Zimbabwe -Joint Research Ethics Committee for the University of Zimbabwe, College of Health Sciences and Parirenyatwa Group of Hospitals REF 221/19).

## Results

Data from 1,052 of 1,101 (96%) children recruited in the PediCAP trial were included in the analysis (**Figure 1**). A small number of children (n=7, 0.7%) were diagnosed with HIV during the trial and were excluded from this analysis. Eleven (1.0%) children with missing data on key parameters used in the risk stratification tools, including mid-upper arm circumference (MUAC), wheezing, and weight-for-height z-score (WHZ), were also excluded; there were no deaths recorded among these 11 children. The median age was 13 months (IQR: 6-23 months; **Table 1**), and 44% of children were female. Most (94%) children had chest indrawing at presentation, 43% had lethargy/unconsciousness, 38% had inability to feed, 24% had wheeze, and 5% had convulsions. The median weight-for-age z-score (WAZ) was - 0.8 (IQR: -1.8-0.0), and the median MUAC was 14cm (IQR: 13-15cm) (**Table 1**).

**Table 1.** Baseline characteristics, overall and split by whether the child died in hospital. Continuous variables show median (IQR).

| variable | Overall (N=1,052) | Death in hospital (N=18) | Discharged alive (N=1,034) |
| --- | --- | --- | --- |
| Female | 458 (43.5%) | 6 (33.3%) | 452 (43.7%) |
| Age (months) | 12.8 (6.3-23.4) | 5.4 (4.1-11.8) | 12.9 (6.4-23.5) |
| Temperature (°C) | 36.8 (36.5-37.6) | 37.9 (37.1-38.5) | 36.8 (36.5-37.5) |
| Respiratory Rate (breaths/min) | 52.0 (42.0-59.0) | 61.0 (52.8-71.5) | 52.0 (42.0-59.0) |
| On Oxygen When Saturation Measured | 274 (26.0%) | 4 (22.2%) | 270 (26.1%) |
| Oxygen Saturation (%) | 95.0 (88.0-98.0) | 80.0 (70.5-88.0) | 95.0 (88.0-98.0) |
| Chest Indrawing | 991 (94.2%) | 18 (100.0%) | 973 (94.1%) |
| Lethargy / unconscious | 450 (42.8%) | 10 (55.6%) | 440 (42.6%) |
| Convulsions | 53 (5.0%) | 3 (16.7%) | 50 (4.8%) |
| Cyanosis | 30 (2.9%) | 1 (5.6%) | 29 (2.8%) |
| Inability to feed | 394 (37.5%) | 15 (83.3%) | 379 (36.7%) |
| Grunting | 384 (36.5%) | 16 (88.9%) | 368 (35.6%) |
| Wheeze | 252 (24.0%) | 0 (0%) | 252 (24.4%) |
| Weight-for-age z-score | -0.8 (-1.8-0.0) | -2.1 (-3.7--1.3) | -0.8 (-1.8-0.1) |
| Weight-for-height z-score | -0.3 (-1.5-0.8) | -1.7 (-3.0--0.2) | -0.3 (-1.5-0.8) |
| Middle upper arm circumference (cm) | 14.0 (13.0-15.1) | 12.6 (10.8-13.0) | 14.0 (13.0-15.2) |
| Centre: South Africa 1 | 236 (22.4%) | 0 (0%) | 236 (22.8%) |
| Centre: Uganda 1 | 191 (18.2%) | 0 (0%) | 191 (18.5%) |
| Centre: Zambia 1 | 157 (14.9%) | 12 (66.7%) | 145 (14%) |
| Centre: Zimbabwe 1 | 141 (13.4%) | 2 (11.1%) | 139 (13.4%) |
| Centre: Uganda 2 | 91 (8.7%) | 3 (16.7%) | 88 (8.5%) |
| Centre: Mozambique 2 | 71 (6.7%) | 0 (0%) | 71 (6.9%) |
| Centre: Mozambique 1 | 55 (5.2%) | 0 (0%) | 55 (5.3%) |
| Centre: Mozambique 4 | 31 (2.9%) | 0 (0%) | 31 (3%) |
| Centre: Mozambique 3 | 27 (2.6%) | 0 (0%) | 27 (2.6%) |
| Centre: Zambia 2 | 24 (2.3%) | 1 (5.6%) | 23 (2.2%) |
| Centre: South Africa 2 | 23 (2.2%) | 0 (0%) | 23 (2.2%) |
| Centre: Zimbabwe 2 | 4 (0.4%) | 0 (0%) | 4 (0.4%) |
| Centre: South Africa 3 | 1 (0.1%) | 0 (0%) | 1 (0.1%) |

**Figure 1.**
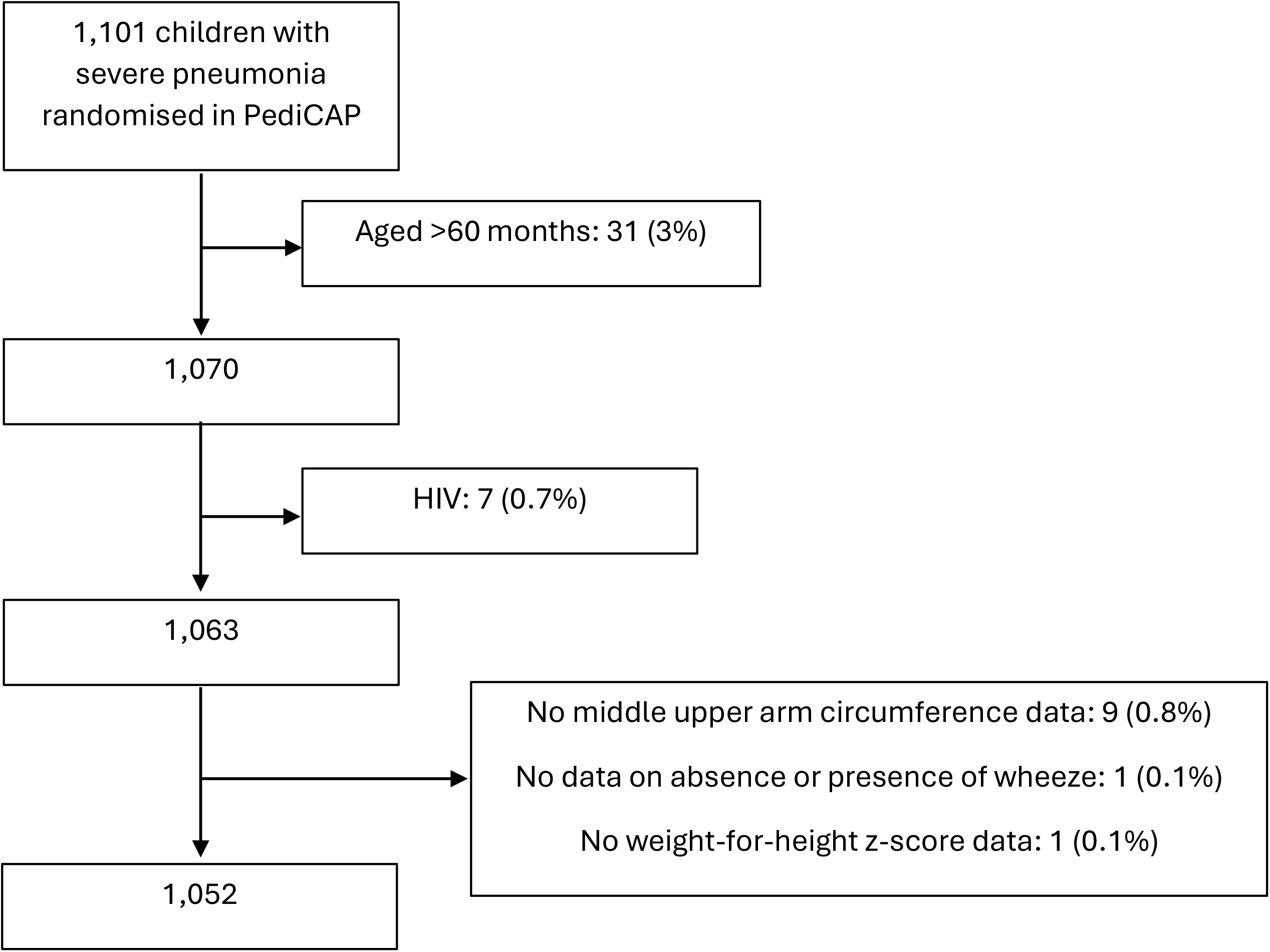
Participant flow from all children randomised in PediCAP to complete cases for all outcomes included in the analysis.

The evaluated scores included between 4 and 10 components (Table S2), with measures of nutritional status, respiratory compromise and neurological status commonly represented. The same components in different scores always agreed on the direction of risk (e.g., females having a higher risk than males).

Applying the various risk scores to the children in PediCAP, the number of distinct score values ranged from 4 in WHO danger signs to 26 in RISC-Malawi (**Table 2**; **Figure S1**), with the percentage of children in the score category representing the greatest proportion of children ranging from 12% in PERCH to 46% in WHO danger signs (**Table 2**). Scores for lethargy/unconscious, chest indrawing, and oxygen saturation were generally important contributors to the total score, though this varied somewhat depending on the points assigned (**Figure S2**).

**Table 2.** Summary of scores points from PediCAP data. Scores are presented in order of publication from newest to oldest, followed by WHO signs. “no unconsciousness” indicates the PediCAP variable ‘lethargy or unconsciousness’ was considered as ‘lethargy’, rather than as ‘unconsciousness’ as in the primary analysis.

| Score | Median (IQR) | Range | Number of distinct score values | Most common score N (%) [score value] |
| --- | --- | --- | --- | --- |
| PREPARE | 3 (1, 4) | 0 - 10 | 11 | 206 (20%) [2] |
| PREPARE: no unconsciousness | 2 (1, 4) | 0 - 10 | 11 | 219 (21%) [2] |
| RISC-KGMU | 2 (2, 3) | -2 - 6 | 8 | 304 (29%) [2] |
| PERCH | 6 (3, 8) | -1 - 16 | 18 | 128 (12%) [6] |
| PERCH: no unconsciousness | 3 (1, 5) | -1 - 11 | 13 | 152 (14%) [1] |
| RISC-Malawi | 8 (1, 11) | -2 - 23 | 26 | 148 (14%) [8] |
| RISC-Malawi: no unconsciousness | 2 (0, 7) | -2 - 15 | 18 | 192 (18%) [0] |
| RISC | 2 (2, 4) | -2 - 6 | 9 | 270 (26%) [2] |
| WHO IMCI pneumonia | 2 (1, 3) | 0 - 5 | 6 | 382 (36%) [2] |
| WHO danger signs | 1 (0, 1) | 0 - 3 | 4 | 476 (45%) [1] |

In-hospital mortality occurred in 18/1052 children; a further 4 children died within 7 days of discharge, giving 22 deaths for the PERCH derivation outcome. Univariate case-fatality rates were highest for variables associated with malnutrition, convulsions, and hypoxaemia (**Figure S3**).

For in-hospital mortality, ROC-AUC point estimates ranged from 0.75 (95% CI:0.28-0.86; WHO danger signs) to 0.87 (95% CI: 0.79-0.95; RISC-KGMU), with point estimates for all scores except WHO danger signs being at least 0.81. Cluster-bootstrap confidence intervals were wide and overlapped substantially across all approaches (**Figure 2**), and no score demonstrated clear superiority. Removing unconsciousness from the models, i.e., treating ‘lethargy or unconsciousness’ as ‘lethargy’ rather than as ‘unconsciousness’ in the primary analysis, led to a small numeric reduction in ROC-AUCs for PREPARE but an increase for PERCH and RISC-Malawi (**Figure 2**). PR-AUC analyses showed similar findings (**Table S3**). Performance at all score thresholds, and optimal threshold based on Youden’s index, is shown in **Table S4**. Sensitivity and specificity varied according to threshold, with no score demonstrating superior performance across thresholds.

**Figure 2.**
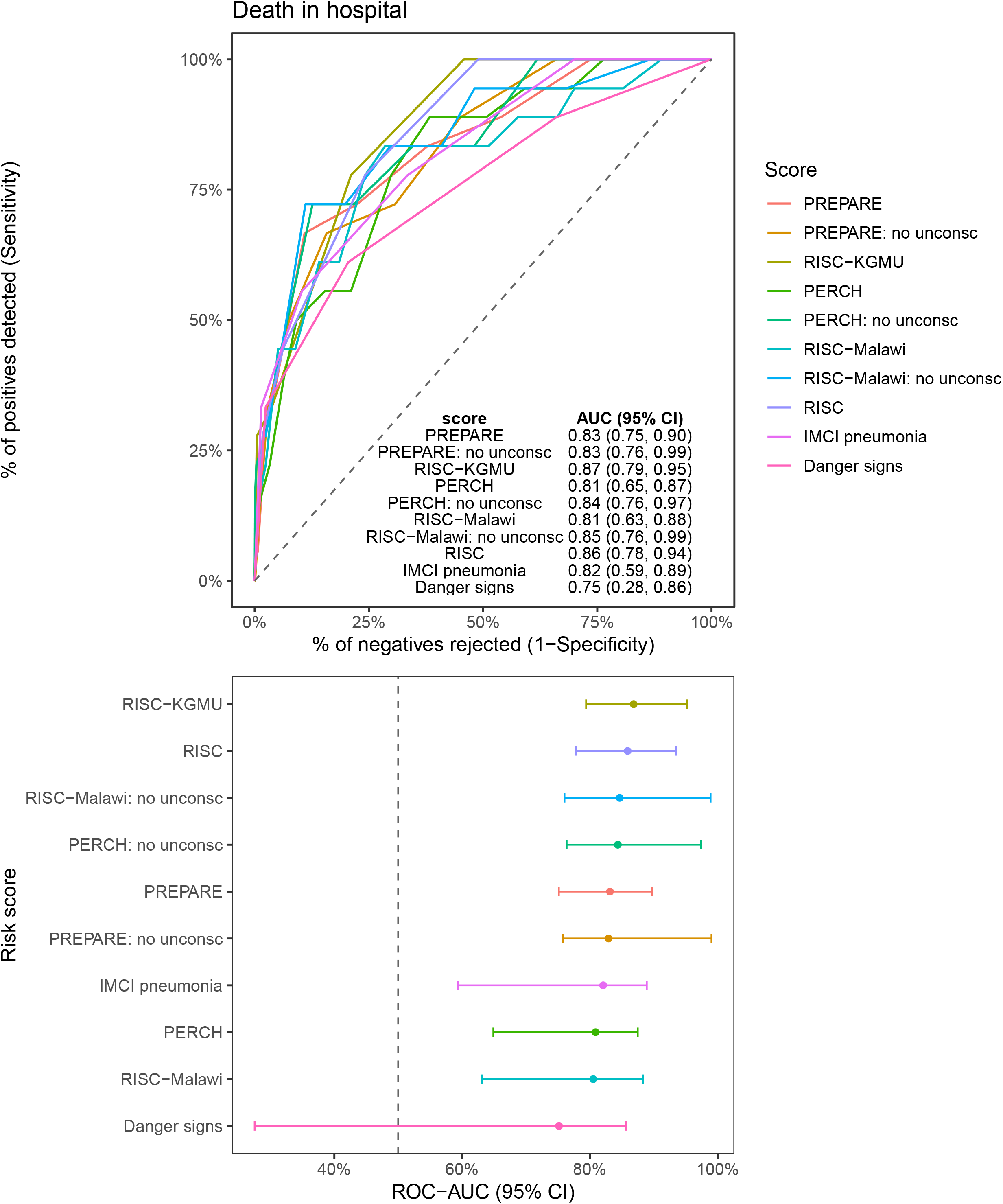
AUC-ROC for each score and the outcome of death in hospital. “no unconsciousness” indicates the PediCAP variable ‘lethargy or unconsciousness’ was considered as ‘lethargy’, rather than as ‘unconsciousness’ as in the primary analysis. The primary outcome for the PERCH risk scores was death in hospital or up to seven days post-discharge; results for this outcome are shown in supplementary figure S4.

Results for mortality during hospitalisation or within 7 days of discharge, the derivation outcome for the PERCH score, were similar to those for mortality during hospitalisation (**Figure S4**). ROC-AUC point estimates ranged from 0.73 to 0.83 and there was substantial overlap in confidence intervals.

Data on readmission or death to day 28 was available for 1010 children. Of these, 63 (6.2%) of children died or were readmitted by day 28 (23 deaths). Exploratory analyses of readmission or death by day 28 demonstrated limited discrimination across all approaches (ROC-AUC 0.54-0.57; **Figure 3**).

**Figure 3.**
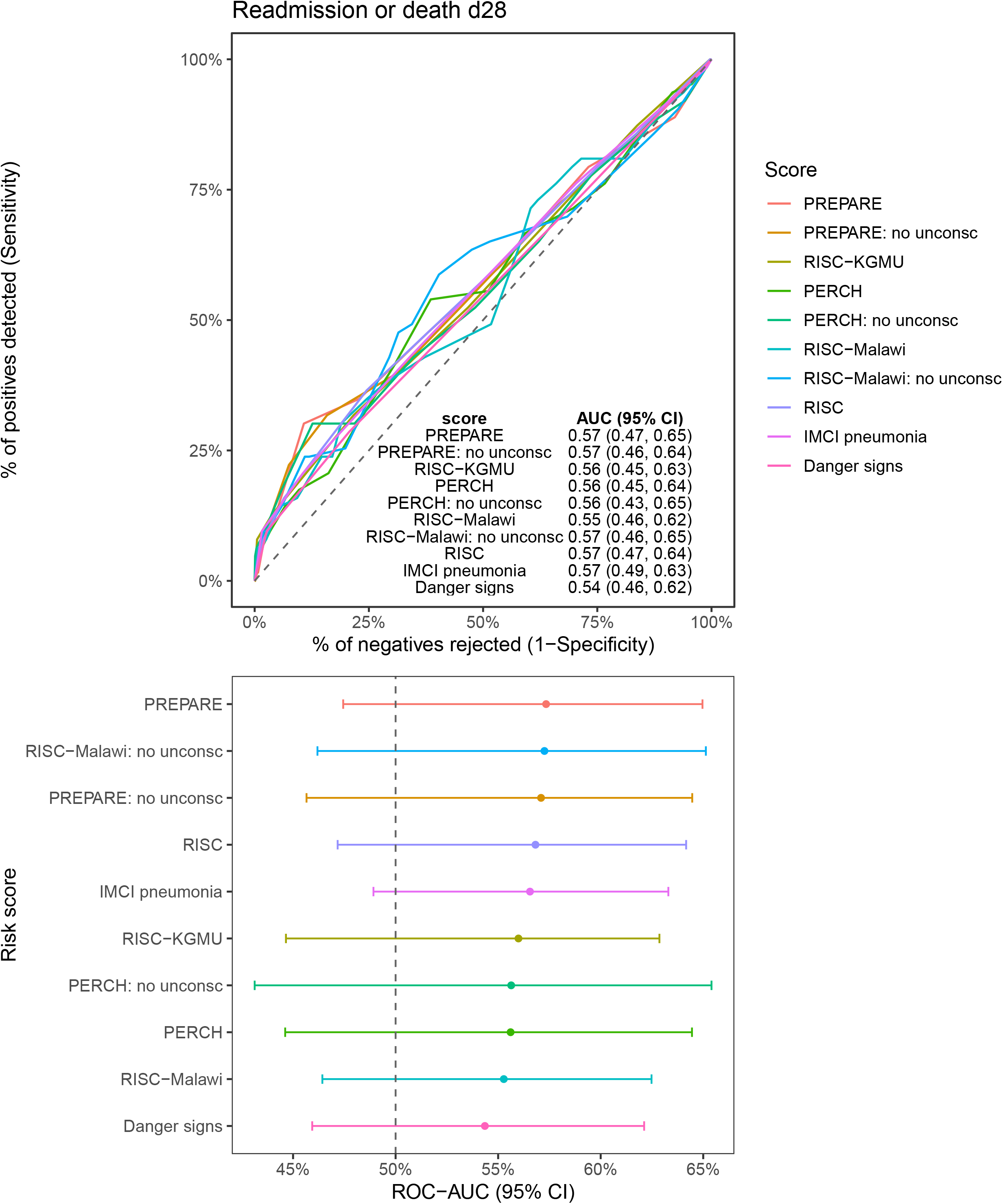
AUC-ROC for each score and the exploratory outcome of readmission or death to day 28. “no unconsciousness” indicates the PediCAP variable ‘lethargy or unconsciousness’ was considered as ‘lethargy’, rather than as ‘unconsciousness’ as in the primary analysis.

## Discussion

Using high-quality, prospectively collected data from children admitted to hospital with severe pneumonia across five African countries within the context of a clinical trial, we evaluated five published risk stratification tools alongside IMCI-based clinical assessment and WHO danger signs. Point estimates for discrimination of mortality outcomes were broadly similar across the published risk scores and WHO IMCI pneumonia criteria (range: 0.81-0.87), whereas WHO danger signs had lower point estimate (0.75). However, confidence intervals were wide owing to the small number of deaths, resulting in considerable uncertainty around relative model performance. Although point estimates for scores other than WHO dangers signs were broadly similar, clinically meaningful differences between approaches remain possible. We found no clear evidence that any single tool consistently outperformed the others for mortality prediction. Strengthening implementation of existing assessment approaches may be as important as introducing additional risk stratification tools.

A key strength of this study is the prospective collection of data across 13 hospitals in five African countries and the direct comparison of multiple published risk scores, WHO IMCI criteria and WHO danger signs within the same population. However, a few limitations should be noted. First, PediCAP was a clinical trial and excluded children living with HIV, those requiring intensive care or invasive ventilation, and those who had already received prolonged intravenous antibiotic treatment. Consequently, our findings may not generalise to these populations, and exclusion of the most severely ill children may have altered the severity spectrum compared with some derivation cohorts. Twelve children were formally excluded from PediCAP because of anticipated need for intensive care or invasive ventilation. However, screening logs were designed to capture children assessed for trial eligibility and were not intended to provide a complete census of all children admitted to intensive care. We therefore cannot reliably estimate the total number of children who may have been excluded on this basis. While exclusion of the most severely ill children may affect the transportability of performance estimates, the direction of this effect is uncertain.

Importantly, all children included in PediCAP had WHO-defined severe pneumonia requiring hospital admission and intravenous antibiotic treatment. Children at the extremes of risk are often more readily identified clinically, whereas the greatest challenge is distinguishing between children at intermediate risk, which is the population represented by much of the PediCAP cohort. Second, we were unable to evaluate risk scores requiring laboratory parameters, as collection of this data was not mandated during the PediCAP trial. Although this prevents comparison with some potentially informative prediction models, laboratory investigations such as blood gas analysis, procalcitonin, and comprehensive blood testing are not routinely available in many resource-constrained settings. Consequently, the clinically based scores evaluated here may be more representative of approaches that could realistically be implemented in routine care. Finally, the relatively small number of mortality events limited precision, meaning clinically meaningful differences between models cannot be excluded. Although complete case analysis ensured comparability across models, some clinical variables are inherently subjective, and differences in measurement variability across sites cannot be ruled out.

Our findings are consistent with most other validation studies for RISC, RISC-Malawi, and PREPARE, which reported ROC-AUC estimates generally within the range observed in our study.^6 11 19^. However, the RISC and PERCH scores performed poorly in a validation study among over 3,000 children aged 0-59 months across 20 LMICs, Australia, and the United States^11^. The difference in performance may reflect differences in case-mix. All children in the PediCAP trial had WHO-defined severe pneumonia, whereas the previous validation included children with pneumonia of varying severity. Risk factors such as hypoxaemia, malnutrition, reduced consciousness, and convulsions may have been more prevalent and potentially more informative for risk stratification in the PediCAP population. WHO IMCI pneumonia criteria generally showed higher point estimates than WHO danger signs, although confidence intervals overlapped substantially, suggesting that inclusion of additional markers of disease severity, such as chest indrawing and oxygen requirement, may modestly improve risk stratification. Taken together, these results show that the risk stratification tools we considered were indeed useful for identifying patients at high risk of death in this population, who require close monitoring from admission throughout hospitalisation. Results were very similar between in-hospital mortality and mortality in-hospital plus 7 days post-discharge, likely because only four of the 22 deaths occurred after discharge. In contrast, all tools performed poorly at predicting the exploratory outcome of 28-day readmission or death. This may be because the scores were designed to predict mortality rather than post-discharge outcomes and do not capture broader social and health system factors influencing readmission^20^. Nevertheless, this outcome remains clinically and programmatically important, as post-discharge events contribute substantially to the overall burden of childhood pneumonia in low-resource settings. The findings also suggest that mortality and readmission reflect different underlying processes. While mortality is more closely related to disease severity and physiological compromise, readmission may additionally be influenced by factors such as access to care, caregiver resources, health-seeking behaviour, and local health-system practices. Composite outcomes combining death and readmission should therefore be selected and interpreted carefully in clinical trials. In PediCAP, this outcome was appropriate because an important concern was that earlier transition to oral antibiotics might lead to treatment failure after discharge requiring readmission. However, such outcomes may be less suitable when the primary objective is assessment of clinical resolution, as readmission may be influenced by factors only partly related to the child’s clinical status.

The variables most strongly associated with in-hospital mortality were related to malnutrition, convulsions and hypoxaemia. Malnutrition is strongly associated with severe adverse outcomes in children with pneumonia^21 22^, likely reflecting the well-described “vicious cycle” between malnutrition and infections. Malnutrition not only increases susceptibility to infections such as pneumonia through immune compromise, but may also increase the severity of infection and risk of death^23^. Routine nutritional assessment is therefore critical in identifying high-risk children who need to be prioritised for close inpatient monitoring and post-discharge follow-up. Convulsions and hypoxaemia were also influential predictors, likely because they are markers of severe illness.

No score demonstrated clear superiority in this study. In the absence of strong evidence favouring one approach, practical considerations such as ease of implementation, familiarity, and availability of required measurements may influence the choice of tool in routine care. Tools that rely on easily collected variables and straightforward assessments, and that do not require calibrated equipment such as weighing scales or stadiometers or the use of WHO z-scores, are more likely to be implemented consistently across health facilities. In this context, scores that use MUAC instead of weight-for-height (e.g., RISC Malawi) may be more practical in some LMIC settings. While WHO danger signs are the simplest and most familiar, their lower point estimates for discrimination should be interpreted cautiously given the limited number of mortality events. Overall, practical considerations including simplicity, familiarity, and measurement requirements are likely to be important when selecting tools for implementation in routine care.

Several important questions remain. Although WHO IMCI criteria and the published risk scores showed broadly similar performance in this study, the limited number of mortality events meant that clinically meaningful differences could not be excluded. Larger datasets or pooled analyses comparing WHO IMCI criteria and published risk scores may help clarify whether any of the published risk scores provide clinically meaningful improvements in discrimination beyond existing WHO criteria. Future work should also evaluate the implementation of risk stratification tools in routine care and determine whether their use improves patient outcomes. Finally, additional research is needed to develop and validate tools for post-discharge outcomes, as readmission and mortality appear to reflect different underlying processes and were poorly captured by existing mortality-focused scores.

## Data Availability

All data produced in the present study are available upon reasonable request to the authors

## Acknowledgements

We would like to thank all the children and caregivers who took part in PediCAP, the clinical teams at participating hospitals, the trial coordination and data management teams, and all members of the PediCAP study team who contributed to the design, conduct, and delivery of the trial.

## Funding

PediCAP was part of the EDCTP2 Programme supported by the European Union (grant number RIA2017MC-2023) and supported all authors. MC and ASW were additionally supported by core support to the MRC Clinical Trials Unit at UCL [MC_UU_00004/04, MC_UU_00004/05]. ASW is a National Institutes of Health and Care Research Senior Investigator. DPM was partially funded by a grant awarded by the Carnegie Corporation of New York.

## Competing interests

Sandoz provided oral amoxicillin and co-amoxiclav fixed dose combination dispersible tablets but had no role in the design, conductor analysis of the trial. The amoxicillin 200 mg/clavulanic acid 28.5 mg 7/1 novel dispersible tablet used in the trial was included in the 2023 WHO Essential Medicines List for Children following a submission from Sandoz. MS Chaired the 2023 WHO Expert Committee on Selection and Use of Essential Medicines. All other authors have no completing interest to declare.

## Author contributions

DPM, ASW, MS and MC contributed to the conception and design of the study. DN contributed to the methodology and investigation, while MC conducted the formal analyses and data curation. DN and MC wrote the first draft of the manuscript. VM, VMu, HAM, MSid, WCB and SAM contributed to data acquisition and investigation. DPM, ASW and MS provided supervision and JAB, ASW and MS secured funding for the study. Members of the PediCAP Study Group contributed to participant recruitment, data collection and study conduct. All authors contributed to interpretation of the findings, critically revised the manuscript for important intellectual content, and approved the final version. MC had full access to all study data, performed the statistical analyses, and takes responsibility for the integrity of the data and accuracy of the analyses as guarantor. Artificial intelligence (co-pilot) was used to improve the scientific writing.

## Data availability

Data is available on reasonable request. Access to PediCAP trial data is subject to approval by the PediCAP Trial Management Group and relevant ethics and governance requirements.

## Notes

### Clinical Trial

ISRCTN63115131

### Author Declarations

Ethical approval was not required for this secondary analysis. For the main PediCAP trial, local ethical approval was sought and obtained in each country. We also obtained hospital or Provincial Health Authority approvals, in addition to regulatory approvals for the clinical trial (Mozambique: Comite Nacional de Bioethica Para a Saude (IRB00002657) REF 323/CNBS/22, South Africa: University of the Witwatersrand Human Research Ethics Committee (Medical) REF 190913B, Uganda: Makarere University Research Ethics Committee REF 2019-162, Zambia: University of Zambia Biomedical Research Ethics Committee REF 328-2019, Zimbabwe: Joint Research Ethics Committee for the University of Zimbabwe, College of Health Sciences and Parirenyatwa Group of Hospitals REF 221/19).

### Summary of Updates

Minor updates following reviewer comments

