## Supplementary information for "Performance of five risk stratification tools for paediatric pneumonia against WHO scores using data from the PediCAP trial in sub-Saharan Africa"

### PediCAP risk scores supplementary information

#### Table S1

Scores from Rees et al 2025^1^ not included in comparison

| # | Paper | Reason not suitable |
| --- | --- | --- |
| 1 | McCollum ED, et al. PLoS One. 2019^2^ | Outpatient score. Water source and sibling data not available in PediCAP. |
| 2 | King C, et al. PLoS One. 2015^3^ | Outpatient score. Number of pneumococcal conjugate vaccine (PCV) doses not available in PediCAP. |
| 3 | Xu, C, et al. Front Pediatr. 2023^4^ | Albumin, haemoglobin, blood pH data not available in PediCAP. |
| 4 | Anteneh ZA, et al. PLoS One. 2023^5^ | Full blood count data not available in PediCAP. |
| 5 | Valentania V, et al. Multidiscip Resp Med. 2021^6^ | Procalcitonin data not available in PediCAP. |
| 6 | Emukule GO, et al. PLoS One. 2014^7^ | Dehydration, night sweats and alert-voice-pain-unconscious (AVPU) scale not available in PediCAP. |
| 7 | Reed C, et al. PLoS One. 2012^8^ (The Respiratory Index of Severity in Children score for HIV-positive children) | HIV positive children mainly excluded from PediCAP due to exclusion criteria of being on long-term antibiotics for prophylaxis. |
| 8 | Mamun et al 2023. PLoS Global Public Health^9^ | Model specification was not included in the publication, and so could not be coded using PediCAP data. |

#### Table S2

Components and associated risk scores included in our analysis. Component names in brackets correspond labels in supplementary figure 1.

| **Component** | **PREPARE**  (outcome: death in hospital) | **RISC-KGMU**  (outcome: death in hospital) | **PERCH**  (outcome: death in hospital + 7 days) | **RISC-Malawi**  (outcome: death in hospital) | **RISC**  (outcome: death in hospital) | **IMCI pneum-onia** | **WHO danger signs** | **N scores** |
| --- | --- | --- | --- | --- | --- | --- | --- | --- |
| **Age** (age) | 2-<6 months: 2 6 -<12 mths:1 ≥12 months: 0 |  | 1-<12 months: 2 ≥12 months: 0 |  |  |  |  | **2** |
| **Sex** (sex) | female: 1 male: 0 |  | female: 1 male: 0 | female: 1 male: 0 |  |  |  | **3** |
| **Weight-for-age z-score** (wfaz) | <-3: 3 -3 to -2: 2 ≥-2: 0 |  |  |  | <-3: 2 -3 to -2: 1 ≥-2: 0 |  |  | **3** |
| **Weight-for-height z-score** (wfhz) |  | <-3: 2 -3 to -2: 1 ≥-2: 0 | <-3: 3 -3 to -2: 2 ≥-2: 0 |  |  | <-3: 1 ≥-3: 0 | <-3: 1 ≥-3: 0 | **3** |
| **Middle upper arm circumference** (muac) |  |  |  | <11.5: 7 11.5 - < 13.5: 3 ≥ 13.5: 0 |  |  |  | **1** |
| **Duration of illness** (max dur) |  |  | <3 days: 0 ≥3 days: 2 |  |  |  |  | **1** |
| **Cyanosis** (cyanosis) | yes: 2 no: 0 |  |  |  |  |  |  | **1** |
| **Oxygen saturation** (o2sat) | <90%: 2 ≥90%: 0 | <90%: 3 ≥90% & chest indrawing: 2 ≥90%, no chest indrawing: 0 | <92%: 2 ≥92%: 0 | <90%: 7 90 - <93%: 2 ≥90%: 0 | <90%: 3 ≥90% & chest indrawing: 2 ≥90%, no chest indrawing: 0 | <90%: 1 ≥90%: 0 |  | **6** |
| **Chest indrawing** (chest indraw) |  |  | Unconscious with chest indrawing: 5 Unconscious, no chest indrawing: 2 Not unconscious: 0 |  |  | yes: 1 no: 0 |  | **5** |
| **Unconscious** (unconscious) | yes: 1 no: 0 |  |  | yes: 8 no: 0 |  | yes: 1 no: 0  (including lethargy) | yes: 1 no: 0  (including lethargy) | **5** |
| **Refusing feed**  (not feeding) |  | yes: 1 no: 0 |  |  | yes: 1 no: 0 | yes: 1 no: 0 | yes: 1 no: 0 | **4** |
| **Convulsions** (convulsions) | yes: 2 no: 0 |  |  |  |  | yes: 1 no: 0 | yes: 1 no: 0 | **3** |
| **Respiratory rate**  (resp rate) | >70 & <12 months:1 >60 & ≥ 12 months: 1 otherwise: 0 |  |  |  |  |  |  | **1** |
| **Temperature** (temp) | <35.5 °C: 3 ≥35.5 °C: 0 |  |  |  |  |  |  | **1** |
| **Cough** (cough) |  |  | yes: -1 no: 0 |  |  |  |  | **1** |
| **Grunting** (grunting) |  |  | yes: 2 no: 0 |  |  |  |  | **1** |
| **Wheeze** (wheeze) |  | yes: -2 no: 0 |  | yes: -2 no: 0 | yes: -2 no: 0 |  |  | **3** |
| **N factors** | **10** | **5** | **9** | **5** | **5** | **6** | **4** |  |
| **Minimum points** | **0** | **-2** | **-1** | **-2** | **-2** | **0** | **0** |  |
| **Maximum points** | **17** | **8** | **17** | **23** | **7** | **6** | **4** |  |
| **Range of points** | **17** | **10** | **18** | **25** | **9** | **6** | **4** |  |

#### Table S3

Precision-recall estimates and 95% confidence intervals for all scores and outcomes

| score | Death in hospital | Death in hospital + 7 days | Readmission or death to d28 |
| --- | --- | --- | --- |
| PREPARE | 0.10 (0.01 - 0.20) | 0.12 (0.02 - 0.19) | 0.10 (0.05 - 0.16) |
| PREPARE: no unconsc | 0.10 (0.02 - 0.18) | 0.12 (0.02 - 0.19) | 0.10 (0.05 - 0.16) |
| RISC-KGMU | 0.20 (0.01 - 0.27) | 0.17 (0.01 - 0.25) | 0.12 (0.04 - 0.20) |
| PERCH | 0.12 (0.01 - 0.21) | 0.11 (0.01 - 0.22) | 0.10 (0.05 - 0.17) |
| PERCH: no unconsc | 0.23 (0.02 - 0.33) | 0.21 (0.02 - 0.33) | 0.14 (0.05 - 0.24) |
| RISC-Malawi | 0.11 (0.00 - 0.34) | 0.14 (0.01 - 0.26) | 0.10 (0.05 - 0.17) |
| RISC-Malawi: no unconsc | 0.12 (0.03 - 0.32) | 0.14 (0.02 - 0.27) | 0.10 (0.05 - 0.16) |
| RISC | 0.12 (0.01 - 0.18) | 0.12 (0.02 - 0.18) | 0.10 (0.05 - 0.14) |
| IMCI pneumonia | 0.14 (0.00 - 0.22) | 0.13 (0.01 - 0.22) | 0.10 (0.05 - 0.17) |
| Danger signs | 0.10 (0.00 - 0.20) | 0.09 (0.01 - 0.20) | 0.09 (0.04 - 0.15) |

#### Table S4

Performance metrics for each score and threshold for the outcome of death in hospital. Cutoffs maximising Youden's index for each score are highted in gray

| **score** | **Score threshold** | **N above threshold** | **% above thres-hold** | **Number of outcomes at thres-hold** | **Number of outcomes above thres-hold** | **Sensitivity (95% CI)** | **Specificity (95% CI)** | **Positive predictive value (95% CI)** | **Negative predictive value (95% CI)** | **Positive likelihood ratio (95% CI)** | **Negative likelihood ratio (95% CI)** |
| --- | --- | --- | --- | --- | --- | --- | --- | --- | --- | --- | --- |
| PREPARE | 10 | 5 | 0.5% | 1 | 1 | 0.06 (0.00, 0.27) | 1.00 (0.99, 1.00) | 0.20 (0.01, 0.72) | 0.98 (0.97, 0.99) | 14.36 (1.69, 122.21) | 0.95 (0.85, 1.06) |
| PREPARE | 9 | 8 | 0.8% | 0 | 1 | 0.06 (0.00, 0.27) | 0.99 (0.99, 1.00) | 0.12 (0.00, 0.53) | 0.98 (0.97, 0.99) | 8.21 (1.06, 63.29) | 0.95 (0.85, 1.06) |
| PREPARE | 8 | 29 | 2.8% | 4 | 5 | 0.28 (0.10, 0.53) | 0.98 (0.97, 0.99) | 0.17 (0.06, 0.36) | 0.99 (0.98, 0.99) | 11.97 (5.15, 27.81) | 0.74 (0.56, 0.98) |
| PREPARE | 7 | 58 | 5.5% | 2 | 7 | 0.39 (0.17, 0.64) | 0.95 (0.94, 0.96) | 0.12 (0.05, 0.23) | 0.99 (0.98, 0.99) | 7.88 (4.17, 14.92) | 0.64 (0.44, 0.93) |
| PREPARE | 6 | 125 | 11.9% | 5 | 12 | 0.67 (0.41, 0.87) | 0.89 (0.87, 0.91) | 0.10 (0.05, 0.16) | 0.99 (0.99, 1.00) | 6.10 (4.21, 8.83) | 0.37 (0.19, 0.72) |
| PREPARE | 5 | 245 | 23.3% | 1 | 13 | 0.72 (0.47, 0.90) | 0.78 (0.75, 0.80) | 0.05 (0.03, 0.09) | 0.99 (0.99, 1.00) | 3.22 (2.37, 4.38) | 0.36 (0.17, 0.75) |
| PREPARE | 4 | 406 | 38.6% | 2 | 15 | 0.83 (0.59, 0.96) | 0.62 (0.59, 0.65) | 0.04 (0.02, 0.06) | 1.00 (0.99, 1.00) | 2.20 (1.77, 2.75) | 0.27 (0.10, 0.75) |
| PREPARE | 3 | 573 | 54.5% | 1 | 16 | 0.89 (0.65, 0.99) | 0.46 (0.43, 0.49) | 0.03 (0.02, 0.04) | 1.00 (0.98, 1.00) | 1.65 (1.39, 1.96) | 0.24 (0.07, 0.89) |
| PREPARE | 2 | 779 | 74.0% | 2 | 18 | 1.00 (0.81, 1.00) | 0.26 (0.24, 0.29) | 0.02 (0.01, 0.04) | 1.00 (0.99, 1.00) | 1.36 | 0 |
| PREPARE | 1 | 969 | 92.1% | 0 | 18 | 1.00 (0.81, 1.00) | 0.08 (0.06, 0.10) | 0.02 (0.01, 0.03) | 1.00 (0.96, 1.00) | 1.09 | 0 |
| PREPARE | 0 | 1052 | 100.0% | 0 | 18 | 1.00 (0.81, 1.00) | 0.00 (0.00, 0.00) | 0.02 (0.01, 0.03) |  | 1 |  |
| PREPARE: no unconsc | 10 | 1 | 0.1% | 0 | 0 | 0.00 (0.00, 0.19) | 1.00 (0.99, 1.00) | 0.00 (0.00, 0.97) | 0.98 (0.97, 0.99) | 0 | 1 |
| PREPARE: no unconsc | 9 | 6 | 0.6% | 1 | 1 | 0.06 (0.00, 0.27) | 1.00 (0.99, 1.00) | 0.17 (0.00, 0.64) | 0.98 (0.97, 0.99) | 11.49 (1.41, 93.44) | 0.95 (0.85, 1.06) |
| PREPARE: no unconsc | 8 | 17 | 1.6% | 2 | 3 | 0.17 (0.04, 0.41) | 0.99 (0.98, 0.99) | 0.18 (0.04, 0.43) | 0.99 (0.98, 0.99) | 12.31 (3.87, 39.13) | 0.84 (0.69, 1.04) |
| PREPARE: no unconsc | 7 | 36 | 3.4% | 3 | 6 | 0.33 (0.13, 0.59) | 0.97 (0.96, 0.98) | 0.17 (0.06, 0.33) | 0.99 (0.98, 0.99) | 11.49 (5.47, 24.14) | 0.69 (0.50, 0.95) |
| PREPARE: no unconsc | 6 | 87 | 8.3% | 3 | 9 | 0.50 (0.26, 0.74) | 0.92 (0.91, 0.94) | 0.10 (0.05, 0.19) | 0.99 (0.98, 1.00) | 6.63 (3.98, 11.03) | 0.54 (0.34, 0.86) |
| PREPARE: no unconsc | 5 | 175 | 16.6% | 3 | 12 | 0.67 (0.41, 0.87) | 0.84 (0.82, 0.86) | 0.07 (0.04, 0.12) | 0.99 (0.99, 1.00) | 4.23 (2.96, 6.04) | 0.40 (0.21, 0.76) |
| PREPARE: no unconsc | 4 | 331 | 31.5% | 1 | 13 | 0.72 (0.47, 0.90) | 0.69 (0.66, 0.72) | 0.04 (0.02, 0.07) | 0.99 (0.98, 1.00) | 2.35 (1.74, 3.17) | 0.40 (0.19, 0.85) |
| PREPARE: no unconsc | 3 | 482 | 45.8% | 3 | 16 | 0.89 (0.65, 0.99) | 0.55 (0.52, 0.58) | 0.03 (0.02, 0.05) | 1.00 (0.99, 1.00) | 1.97 (1.65, 2.35) | 0.20 (0.05, 0.75) |
| PREPARE: no unconsc | 2 | 701 | 66.6% | 2 | 18 | 1.00 (0.81, 1.00) | 0.34 (0.31, 0.37) | 0.03 (0.02, 0.04) | 1.00 (0.99, 1.00) | 1.51 | 0 |
| PREPARE: no unconsc | 1 | 911 | 86.6% | 0 | 18 | 1.00 (0.81, 1.00) | 0.14 (0.12, 0.16) | 0.02 (0.01, 0.03) | 1.00 (0.97, 1.00) | 1.16 | 0 |
| PREPARE: no unconsc | 0 | 1052 | 100.0% | 0 | 18 | 1.00 (0.81, 1.00) | 0.00 (0.00, 0.00) | 0.02 (0.01, 0.03) |  | 1 |  |
| RISC-KGMU | 6 | 10 | 1.0% | 5 | 5 | 0.28 (0.10, 0.53) | 1.00 (0.99, 1.00) | 0.50 (0.19, 0.81) | 0.99 (0.98, 0.99) | 57.44 (18.21, 181.18) | 0.73 (0.54, 0.97) |
| RISC-KGMU | 5 | 45 | 4.3% | 1 | 6 | 0.33 (0.13, 0.59) | 0.96 (0.95, 0.97) | 0.13 (0.05, 0.27) | 0.99 (0.98, 0.99) | 8.84 (4.29, 18.20) | 0.69 (0.50, 0.96) |
| RISC-KGMU | 4 | 232 | 22.1% | 8 | 14 | 0.78 (0.52, 0.94) | 0.79 (0.76, 0.81) | 0.06 (0.03, 0.10) | 1.00 (0.99, 1.00) | 3.69 (2.81, 4.85) | 0.28 (0.12, 0.67) |
| RISC-KGMU | 3 | 492 | 46.8% | 4 | 18 | 1.00 (0.81, 1.00) | 0.54 (0.51, 0.57) | 0.04 (0.02, 0.06) | 1.00 (0.99, 1.00) | 2.18 | 0 |
| RISC-KGMU | 2 | 796 | 75.7% | 0 | 18 | 1.00 (0.81, 1.00) | 0.25 (0.22, 0.28) | 0.02 (0.01, 0.04) | 1.00 (0.99, 1.00) | 1.33 | 0 |
| RISC-KGMU | 1 | 883 | 83.9% | 0 | 18 | 1.00 (0.81, 1.00) | 0.16 (0.14, 0.19) | 0.02 (0.01, 0.03) | 1.00 (0.98, 1.00) | 1.2 | 0 |
| RISC-KGMU | 0 | 1048 | 99.6% | 0 | 18 | 1.00 (0.81, 1.00) | 0.00 (0.00, 0.01) | 0.02 (0.01, 0.03) | 1.00 (0.40, 1.00) | 1 | 0 |
| RISC-KGMU | -1 | 1049 | 99.7% | 0 | 18 | 1.00 (0.81, 1.00) | 0.00 (0.00, 0.01) | 0.02 (0.01, 0.03) | 1.00 (0.29, 1.00) | 1 | 0 |
| RISC-KGMU | -2 | 1052 | 100.0% | 0 | 18 | 1.00 (0.81, 1.00) | 0.00 (0.00, 0.00) | 0.02 (0.01, 0.03) |  | 1 |  |
| PERCH | 16 | 2 | 0.2% | 1 | 1 | 0.06 (0.00, 0.27) | 1.00 (0.99, 1.00) | 0.50 (0.01, 0.99) | 0.98 (0.97, 0.99) | 57.44 (3.74, 882.91) | 0.95 (0.85, 1.06) |
| PERCH | 15 | 6 | 0.6% | 1 | 2 | 0.11 (0.01, 0.35) | 1.00 (0.99, 1.00) | 0.33 (0.04, 0.78) | 0.98 (0.98, 0.99) | 28.72 (5.62, 146.92) | 0.89 (0.76, 1.05) |
| PERCH | 14 | 19 | 1.8% | 1 | 3 | 0.17 (0.04, 0.41) | 0.98 (0.97, 0.99) | 0.16 (0.03, 0.40) | 0.99 (0.98, 0.99) | 10.77 (3.44, 33.74) | 0.85 (0.69, 1.04) |
| PERCH | 13 | 38 | 3.6% | 1 | 4 | 0.22 (0.06, 0.48) | 0.97 (0.95, 0.98) | 0.11 (0.03, 0.25) | 0.99 (0.98, 0.99) | 6.76 (2.68, 17.05) | 0.80 (0.63, 1.03) |
| PERCH | 12 | 73 | 6.9% | 3 | 7 | 0.39 (0.17, 0.64) | 0.94 (0.92, 0.95) | 0.10 (0.04, 0.19) | 0.99 (0.98, 0.99) | 6.09 (3.26, 11.38) | 0.65 (0.45, 0.94) |
| PERCH | 11 | 104 | 9.9% | 2 | 9 | 0.50 (0.26, 0.74) | 0.91 (0.89, 0.93) | 0.09 (0.04, 0.16) | 0.99 (0.98, 1.00) | 5.44 (3.30, 8.97) | 0.55 (0.35, 0.87) |
| PERCH | 10 | 169 | 16.1% | 1 | 10 | 0.56 (0.31, 0.78) | 0.85 (0.82, 0.87) | 0.06 (0.03, 0.11) | 0.99 (0.98, 1.00) | 3.61 (2.33, 5.59) | 0.53 (0.31, 0.88) |
| PERCH | 9 | 228 | 21.7% | 0 | 10 | 0.56 (0.31, 0.78) | 0.79 (0.76, 0.81) | 0.04 (0.02, 0.08) | 0.99 (0.98, 1.00) | 2.64 (1.71, 4.05) | 0.56 (0.34, 0.94) |
| PERCH | 8 | 323 | 30.7% | 4 | 14 | 0.78 (0.52, 0.94) | 0.70 (0.67, 0.73) | 0.04 (0.02, 0.07) | 0.99 (0.99, 1.00) | 2.60 (2.00, 3.39) | 0.32 (0.13, 0.75) |
| PERCH | 7 | 412 | 39.2% | 2 | 16 | 0.89 (0.65, 0.99) | 0.62 (0.59, 0.65) | 0.04 (0.02, 0.06) | 1.00 (0.99, 1.00) | 2.32 (1.94, 2.78) | 0.18 (0.05, 0.67) |
| PERCH | 6 | 540 | 51.3% | 0 | 16 | 0.89 (0.65, 0.99) | 0.49 (0.46, 0.52) | 0.03 (0.02, 0.05) | 1.00 (0.99, 1.00) | 1.75 (1.47, 2.09) | 0.23 (0.06, 0.83) |
| PERCH | 5 | 629 | 59.8% | 1 | 17 | 0.94 (0.73, 1.00) | 0.41 (0.38, 0.44) | 0.03 (0.02, 0.04) | 1.00 (0.99, 1.00) | 1.60 (1.41, 1.80) | 0.14 (0.02, 0.92) |
| PERCH | 4 | 734 | 69.8% | 0 | 17 | 0.94 (0.73, 1.00) | 0.31 (0.28, 0.34) | 0.02 (0.01, 0.04) | 1.00 (0.98, 1.00) | 1.36 (1.21, 1.53) | 0.18 (0.03, 1.22) |
| PERCH | 3 | 808 | 76.8% | 1 | 18 | 1.00 (0.81, 1.00) | 0.24 (0.21, 0.26) | 0.02 (0.01, 0.03) | 1.00 (0.98, 1.00) | 1.31 | 0 |
| PERCH | 2 | 878 | 83.5% | 0 | 18 | 1.00 (0.81, 1.00) | 0.17 (0.15, 0.19) | 0.02 (0.01, 0.03) | 1.00 (0.98, 1.00) | 1.2 | 0 |
| PERCH | 1 | 966 | 91.8% | 0 | 18 | 1.00 (0.81, 1.00) | 0.08 (0.07, 0.10) | 0.02 (0.01, 0.03) | 1.00 (0.96, 1.00) | 1.09 | 0 |
| PERCH | 0 | 1008 | 95.8% | 0 | 18 | 1.00 (0.81, 1.00) | 0.04 (0.03, 0.06) | 0.02 (0.01, 0.03) | 1.00 (0.92, 1.00) | 1.04 | 0 |
| PERCH | -1 | 1052 | 100.0% | 0 | 18 | 1.00 (0.81, 1.00) | 0.00 (0.00, 0.00) | 0.02 (0.01, 0.03) |  | 1 |  |
| PERCH: no unconsc | 11 | 4 | 0.4% | 3 | 3 | 0.17 (0.04, 0.41) | 1.00 (0.99, 1.00) | 0.75 (0.19, 0.99) | 0.99 (0.98, 0.99) | 172.33 (18.82, 1578.38) | 0.83 (0.68, 1.03) |
| PERCH: no unconsc | 10 | 8 | 0.8% | 1 | 4 | 0.22 (0.06, 0.48) | 1.00 (0.99, 1.00) | 0.50 (0.16, 0.84) | 0.99 (0.98, 0.99) | 57.44 (15.57, 211.89) | 0.78 (0.61, 1.00) |
| PERCH: no unconsc | 9 | 28 | 2.7% | 1 | 5 | 0.28 (0.10, 0.53) | 0.98 (0.97, 0.99) | 0.18 (0.06, 0.37) | 0.99 (0.98, 0.99) | 12.49 (5.35, 29.14) | 0.74 (0.55, 0.98) |
| PERCH: no unconsc | 8 | 68 | 6.5% | 3 | 8 | 0.44 (0.22, 0.69) | 0.94 (0.93, 0.96) | 0.12 (0.05, 0.22) | 0.99 (0.98, 1.00) | 7.66 (4.32, 13.57) | 0.59 (0.39, 0.89) |
| PERCH: no unconsc | 7 | 144 | 13.7% | 5 | 13 | 0.72 (0.47, 0.90) | 0.87 (0.85, 0.89) | 0.09 (0.05, 0.15) | 0.99 (0.99, 1.00) | 5.70 (4.11, 7.91) | 0.32 (0.15, 0.67) |
| PERCH: no unconsc | 6 | 238 | 22.6% | 0 | 13 | 0.72 (0.47, 0.90) | 0.78 (0.76, 0.81) | 0.05 (0.03, 0.09) | 0.99 (0.99, 1.00) | 3.32 (2.44, 4.52) | 0.36 (0.17, 0.75) |
| PERCH: no unconsc | 5 | 373 | 35.5% | 2 | 15 | 0.83 (0.59, 0.96) | 0.65 (0.62, 0.68) | 0.04 (0.02, 0.07) | 1.00 (0.99, 1.00) | 2.41 (1.93, 3.01) | 0.25 (0.09, 0.72) |
| PERCH: no unconsc | 4 | 512 | 48.7% | 0 | 15 | 0.83 (0.59, 0.96) | 0.52 (0.49, 0.55) | 0.03 (0.02, 0.05) | 0.99 (0.98, 1.00) | 1.73 (1.40, 2.15) | 0.32 (0.11, 0.90) |
| PERCH: no unconsc | 3 | 658 | 62.5% | 3 | 18 | 1.00 (0.81, 1.00) | 0.38 (0.35, 0.41) | 0.03 (0.02, 0.04) | 1.00 (0.99, 1.00) | 1.62 | 0 |
| PERCH: no unconsc | 2 | 781 | 74.2% | 0 | 18 | 1.00 (0.81, 1.00) | 0.26 (0.24, 0.29) | 0.02 (0.01, 0.04) | 1.00 (0.99, 1.00) | 1.36 | 0 |
| PERCH: no unconsc | 1 | 933 | 88.7% | 0 | 18 | 1.00 (0.81, 1.00) | 0.12 (0.10, 0.14) | 0.02 (0.01, 0.03) | 1.00 (0.97, 1.00) | 1.13 | 0 |
| PERCH: no unconsc | 0 | 991 | 94.2% | 0 | 18 | 1.00 (0.81, 1.00) | 0.06 (0.05, 0.08) | 0.02 (0.01, 0.03) | 1.00 (0.94, 1.00) | 1.06 | 0 |
| PERCH: no unconsc | -1 | 1052 | 100.0% | 0 | 18 | 1.00 (0.81, 1.00) | 0.00 (0.00, 0.00) | 0.02 (0.01, 0.03) |  | 1 |  |
| RISC-Malawi | 23 | 5 | 0.5% | 1 | 1 | 0.06 (0.00, 0.27) | 1.00 (0.99, 1.00) | 0.20 (0.01, 0.72) | 0.98 (0.97, 0.99) | 14.36 (1.69, 122.21) | 0.95 (0.85, 1.06) |
| RISC-Malawi | 22 | 11 | 1.0% | 2 | 3 | 0.17 (0.04, 0.41) | 0.99 (0.98, 1.00) | 0.27 (0.06, 0.61) | 0.99 (0.98, 0.99) | 21.54 (6.22, 74.62) | 0.84 (0.68, 1.03) |
| RISC-Malawi | 21 | 14 | 1.3% | 0 | 3 | 0.17 (0.04, 0.41) | 0.99 (0.98, 0.99) | 0.21 (0.05, 0.51) | 0.99 (0.98, 0.99) | 15.67 (4.77, 51.42) | 0.84 (0.69, 1.04) |
| RISC-Malawi | 20 | 15 | 1.4% | 0 | 3 | 0.17 (0.04, 0.41) | 0.99 (0.98, 0.99) | 0.20 (0.04, 0.48) | 0.99 (0.98, 0.99) | 14.36 (4.43, 46.56) | 0.84 (0.69, 1.04) |
| RISC-Malawi | 19 | 29 | 2.8% | 1 | 4 | 0.22 (0.06, 0.48) | 0.98 (0.96, 0.98) | 0.14 (0.04, 0.32) | 0.99 (0.98, 0.99) | 9.19 (3.57, 23.70) | 0.80 (0.62, 1.02) |
| RISC-Malawi | 18 | 61 | 5.8% | 4 | 8 | 0.44 (0.22, 0.69) | 0.95 (0.93, 0.96) | 0.13 (0.06, 0.24) | 0.99 (0.98, 1.00) | 8.67 (4.86, 15.48) | 0.59 (0.39, 0.89) |
| RISC-Malawi | 17 | 63 | 6.0% | 0 | 8 | 0.44 (0.22, 0.69) | 0.95 (0.93, 0.96) | 0.13 (0.06, 0.23) | 0.99 (0.98, 1.00) | 8.36 (4.69, 14.88) | 0.59 (0.39, 0.89) |
| RISC-Malawi | 16 | 100 | 9.5% | 0 | 8 | 0.44 (0.22, 0.69) | 0.91 (0.89, 0.93) | 0.08 (0.04, 0.15) | 0.99 (0.98, 0.99) | 5.00 (2.88, 8.68) | 0.61 (0.40, 0.92) |
| RISC-Malawi | 15 | 157 | 14.9% | 3 | 11 | 0.61 (0.36, 0.83) | 0.86 (0.84, 0.88) | 0.07 (0.04, 0.12) | 0.99 (0.98, 1.00) | 4.33 (2.91, 6.44) | 0.45 (0.25, 0.81) |
| RISC-Malawi | 14 | 166 | 15.8% | 0 | 11 | 0.61 (0.36, 0.83) | 0.85 (0.83, 0.87) | 0.07 (0.03, 0.12) | 0.99 (0.98, 1.00) | 4.08 (2.74, 6.06) | 0.46 (0.26, 0.82) |
| RISC-Malawi | 13 | 180 | 17.1% | 0 | 11 | 0.61 (0.36, 0.83) | 0.84 (0.81, 0.86) | 0.06 (0.03, 0.11) | 0.99 (0.98, 1.00) | 3.74 (2.52, 5.54) | 0.46 (0.26, 0.83) |
| RISC-Malawi | 12 | 202 | 19.2% | 0 | 11 | 0.61 (0.36, 0.83) | 0.82 (0.79, 0.84) | 0.05 (0.03, 0.10) | 0.99 (0.98, 1.00) | 3.31 (2.24, 4.89) | 0.48 (0.27, 0.85) |
| RISC-Malawi | 11 | 264 | 25.1% | 3 | 14 | 0.78 (0.52, 0.94) | 0.76 (0.73, 0.78) | 0.05 (0.03, 0.09) | 0.99 (0.99, 1.00) | 3.22 (2.46, 4.21) | 0.29 (0.12, 0.70) |
| RISC-Malawi | 10 | 310 | 29.5% | 1 | 15 | 0.83 (0.59, 0.96) | 0.71 (0.69, 0.74) | 0.05 (0.03, 0.08) | 1.00 (0.99, 1.00) | 2.92 (2.33, 3.67) | 0.23 (0.08, 0.66) |
| RISC-Malawi | 9 | 396 | 37.6% | 0 | 15 | 0.83 (0.59, 0.96) | 0.63 (0.60, 0.66) | 0.04 (0.02, 0.06) | 1.00 (0.99, 1.00) | 2.26 (1.81, 2.82) | 0.26 (0.09, 0.74) |
| RISC-Malawi | 8 | 544 | 51.7% | 0 | 15 | 0.83 (0.59, 0.96) | 0.49 (0.46, 0.52) | 0.03 (0.02, 0.05) | 0.99 (0.98, 1.00) | 1.63 (1.31, 2.02) | 0.34 (0.12, 0.96) |
| RISC-Malawi | 7 | 612 | 58.2% | 1 | 16 | 0.89 (0.65, 0.99) | 0.42 (0.39, 0.45) | 0.03 (0.02, 0.04) | 1.00 (0.98, 1.00) | 1.54 (1.30, 1.83) | 0.26 (0.07, 0.97) |
| RISC-Malawi | 6 | 643 | 61.1% | 0 | 16 | 0.89 (0.65, 0.99) | 0.39 (0.36, 0.42) | 0.02 (0.01, 0.04) | 1.00 (0.98, 1.00) | 1.47 (1.24, 1.74) | 0.28 (0.08, 1.04) |
| RISC-Malawi | 5 | 660 | 62.7% | 0 | 16 | 0.89 (0.65, 0.99) | 0.38 (0.35, 0.41) | 0.02 (0.01, 0.04) | 0.99 (0.98, 1.00) | 1.43 (1.20, 1.69) | 0.29 (0.08, 1.09) |
| RISC-Malawi | 4 | 701 | 66.6% | 0 | 16 | 0.89 (0.65, 0.99) | 0.34 (0.31, 0.37) | 0.02 (0.01, 0.04) | 0.99 (0.98, 1.00) | 1.34 (1.13, 1.59) | 0.33 (0.09, 1.22) |
| RISC-Malawi | 3 | 741 | 70.4% | 1 | 17 | 0.94 (0.73, 1.00) | 0.30 (0.27, 0.33) | 0.02 (0.01, 0.04) | 1.00 (0.98, 1.00) | 1.35 (1.20, 1.52) | 0.19 (0.03, 1.25) |
| RISC-Malawi | 2 | 761 | 72.3% | 0 | 17 | 0.94 (0.73, 1.00) | 0.28 (0.25, 0.31) | 0.02 (0.01, 0.04) | 1.00 (0.98, 1.00) | 1.31 (1.17, 1.48) | 0.20 (0.03, 1.33) |
| RISC-Malawi | 1 | 851 | 80.9% | 0 | 17 | 0.94 (0.73, 1.00) | 0.19 (0.17, 0.22) | 0.02 (0.01, 0.03) | 1.00 (0.97, 1.00) | 1.17 (1.04, 1.31) | 0.29 (0.04, 1.94) |
| RISC-Malawi | 0 | 939 | 89.3% | 1 | 18 | 1.00 (0.81, 1.00) | 0.11 (0.09, 0.13) | 0.02 (0.01, 0.03) | 1.00 (0.97, 1.00) | 1.12 | 0 |
| RISC-Malawi | -1 | 987 | 93.8% | 0 | 18 | 1.00 (0.81, 1.00) | 0.06 (0.05, 0.08) | 0.02 (0.01, 0.03) | 1.00 (0.94, 1.00) | 1.07 | 0 |
| RISC-Malawi | -2 | 1052 | 100.0% | 0 | 18 | 1.00 (0.81, 1.00) | 0.00 (0.00, 0.00) | 0.02 (0.01, 0.03) |  | 1 |  |
| RISC-Malawi: no unconsc | 15 | 15 | 1.4% | 3 | 3 | 0.17 (0.04, 0.41) | 0.99 (0.98, 0.99) | 0.20 (0.04, 0.48) | 0.99 (0.98, 0.99) | 14.36 (4.43, 46.56) | 0.84 (0.69, 1.04) |
| RISC-Malawi: no unconsc | 14 | 25 | 2.4% | 2 | 5 | 0.28 (0.10, 0.53) | 0.98 (0.97, 0.99) | 0.20 (0.07, 0.41) | 0.99 (0.98, 0.99) | 14.36 (6.06, 34.01) | 0.74 (0.55, 0.98) |
| RISC-Malawi: no unconsc | 13 | 29 | 2.8% | 0 | 5 | 0.28 (0.10, 0.53) | 0.98 (0.97, 0.99) | 0.17 (0.06, 0.36) | 0.99 (0.98, 0.99) | 11.97 (5.15, 27.81) | 0.74 (0.56, 0.98) |
| RISC-Malawi: no unconsc | 12 | 30 | 2.9% | 0 | 5 | 0.28 (0.10, 0.53) | 0.98 (0.96, 0.98) | 0.17 (0.06, 0.35) | 0.99 (0.98, 0.99) | 11.49 (4.96, 26.60) | 0.74 (0.56, 0.99) |
| RISC-Malawi: no unconsc | 11 | 69 | 6.6% | 3 | 8 | 0.44 (0.22, 0.69) | 0.94 (0.92, 0.95) | 0.12 (0.05, 0.22) | 0.99 (0.98, 1.00) | 7.53 (4.26, 13.33) | 0.59 (0.39, 0.89) |
| RISC-Malawi: no unconsc | 10 | 128 | 12.2% | 5 | 13 | 0.72 (0.47, 0.90) | 0.89 (0.87, 0.91) | 0.10 (0.06, 0.17) | 0.99 (0.99, 1.00) | 6.49 (4.65, 9.07) | 0.31 (0.15, 0.66) |
| RISC-Malawi: no unconsc | 9 | 137 | 13.0% | 0 | 13 | 0.72 (0.47, 0.90) | 0.88 (0.86, 0.90) | 0.09 (0.05, 0.16) | 0.99 (0.99, 1.00) | 6.02 (4.33, 8.38) | 0.32 (0.15, 0.67) |
| RISC-Malawi: no unconsc | 8 | 218 | 20.7% | 0 | 13 | 0.72 (0.47, 0.90) | 0.80 (0.78, 0.83) | 0.06 (0.03, 0.10) | 0.99 (0.99, 1.00) | 3.64 (2.67, 4.97) | 0.35 (0.16, 0.73) |
| RISC-Malawi: no unconsc | 7 | 322 | 30.6% | 2 | 15 | 0.83 (0.59, 0.96) | 0.70 (0.67, 0.73) | 0.05 (0.03, 0.08) | 1.00 (0.99, 1.00) | 2.81 (2.24, 3.52) | 0.24 (0.08, 0.67) |
| RISC-Malawi: no unconsc | 6 | 345 | 32.8% | 0 | 15 | 0.83 (0.59, 0.96) | 0.68 (0.65, 0.71) | 0.04 (0.02, 0.07) | 1.00 (0.99, 1.00) | 2.61 (2.09, 3.27) | 0.24 (0.09, 0.69) |
| RISC-Malawi: no unconsc | 5 | 375 | 35.6% | 0 | 15 | 0.83 (0.59, 0.96) | 0.65 (0.62, 0.68) | 0.04 (0.02, 0.07) | 1.00 (0.99, 1.00) | 2.39 (1.92, 2.99) | 0.26 (0.09, 0.72) |
| RISC-Malawi: no unconsc | 4 | 438 | 41.6% | 0 | 15 | 0.83 (0.59, 0.96) | 0.59 (0.56, 0.62) | 0.03 (0.02, 0.06) | 1.00 (0.99, 1.00) | 2.04 (1.64, 2.54) | 0.28 (0.10, 0.79) |
| RISC-Malawi: no unconsc | 3 | 515 | 49.0% | 2 | 17 | 0.94 (0.73, 1.00) | 0.52 (0.49, 0.55) | 0.03 (0.02, 0.05) | 1.00 (0.99, 1.00) | 1.96 (1.72, 2.23) | 0.11 (0.02, 0.72) |
| RISC-Malawi: no unconsc | 2 | 554 | 52.7% | 0 | 17 | 0.94 (0.73, 1.00) | 0.48 (0.45, 0.51) | 0.03 (0.02, 0.05) | 1.00 (0.99, 1.00) | 1.82 (1.60, 2.06) | 0.12 (0.02, 0.78) |
| RISC-Malawi: no unconsc | 1 | 723 | 68.7% | 0 | 17 | 0.94 (0.73, 1.00) | 0.32 (0.29, 0.35) | 0.02 (0.01, 0.04) | 1.00 (0.98, 1.00) | 1.38 (1.23, 1.56) | 0.18 (0.03, 1.18) |
| RISC-Malawi: no unconsc | 0 | 915 | 87.0% | 1 | 18 | 1.00 (0.81, 1.00) | 0.13 (0.11, 0.15) | 0.02 (0.01, 0.03) | 1.00 (0.97, 1.00) | 1.15 | 0 |
| RISC-Malawi: no unconsc | -1 | 974 | 92.6% | 0 | 18 | 1.00 (0.81, 1.00) | 0.08 (0.06, 0.09) | 0.02 (0.01, 0.03) | 1.00 (0.95, 1.00) | 1.08 | 0 |
| RISC-Malawi: no unconsc | -2 | 1052 | 100.0% | 0 | 18 | 1.00 (0.81, 1.00) | 0.00 (0.00, 0.00) | 0.02 (0.01, 0.03) |  | 1 |  |
| RISC | 6 | 23 | 2.2% | 5 | 5 | 0.28 (0.10, 0.53) | 0.98 (0.97, 0.99) | 0.22 (0.07, 0.44) | 0.99 (0.98, 0.99) | 15.96 (6.66, 38.26) | 0.74 (0.55, 0.98) |
| RISC | 5 | 72 | 6.8% | 3 | 8 | 0.44 (0.22, 0.69) | 0.94 (0.92, 0.95) | 0.11 (0.05, 0.21) | 0.99 (0.98, 1.00) | 7.18 (4.07, 12.68) | 0.59 (0.39, 0.90) |
| RISC | 4 | 264 | 25.1% | 6 | 14 | 0.78 (0.52, 0.94) | 0.76 (0.73, 0.78) | 0.05 (0.03, 0.09) | 0.99 (0.99, 1.00) | 3.22 (2.46, 4.21) | 0.29 (0.12, 0.70) |
| RISC | 3 | 524 | 49.8% | 4 | 18 | 1.00 (0.81, 1.00) | 0.51 (0.48, 0.54) | 0.03 (0.02, 0.05) | 1.00 (0.99, 1.00) | 2.04 | 0 |
| RISC | 2 | 794 | 75.5% | 0 | 18 | 1.00 (0.81, 1.00) | 0.25 (0.22, 0.28) | 0.02 (0.01, 0.04) | 1.00 (0.99, 1.00) | 1.33 | 0 |
| RISC | 1 | 881 | 83.7% | 0 | 18 | 1.00 (0.81, 1.00) | 0.17 (0.14, 0.19) | 0.02 (0.01, 0.03) | 1.00 (0.98, 1.00) | 1.2 | 0 |
| RISC | 0 | 1048 | 99.6% | 0 | 18 | 1.00 (0.81, 1.00) | 0.00 (0.00, 0.01) | 0.02 (0.01, 0.03) | 1.00 (0.40, 1.00) | 1 | 0 |
| RISC | -1 | 1050 | 99.8% | 0 | 18 | 1.00 (0.81, 1.00) | 0.00 (0.00, 0.01) | 0.02 (0.01, 0.03) | 1.00 (0.16, 1.00) | 1 | 0 |
| RISC | -2 | 1052 | 100.0% | 0 | 18 | 1.00 (0.81, 1.00) | 0.00 (0.00, 0.00) | 0.02 (0.01, 0.03) |  | 1 |  |
| IMCI pneumonia | 5 | 21 | 2.0% | 6 | 6 | 0.33 (0.13, 0.59) | 0.99 (0.98, 0.99) | 0.29 (0.11, 0.52) | 0.99 (0.98, 0.99) | 22.98 (10.08, 52.39) | 0.68 (0.49, 0.94) |
| IMCI pneumonia | 4 | 117 | 11.1% | 4 | 10 | 0.56 (0.31, 0.78) | 0.90 (0.88, 0.91) | 0.09 (0.04, 0.15) | 0.99 (0.98, 1.00) | 5.37 (3.42, 8.42) | 0.50 (0.30, 0.83) |
| IMCI pneumonia | 3 | 360 | 34.2% | 4 | 14 | 0.78 (0.52, 0.94) | 0.67 (0.64, 0.69) | 0.04 (0.02, 0.06) | 0.99 (0.99, 1.00) | 2.32 (1.79, 3.02) | 0.33 (0.14, 0.79) |
| IMCI pneumonia | 2 | 742 | 70.5% | 4 | 18 | 1.00 (0.81, 1.00) | 0.30 (0.27, 0.33) | 0.02 (0.01, 0.04) | 1.00 (0.99, 1.00) | 1.43 | 0 |
| IMCI pneumonia | 1 | 1040 | 98.9% | 0 | 18 | 1.00 (0.81, 1.00) | 0.01 (0.01, 0.02) | 0.02 (0.01, 0.03) | 1.00 (0.74, 1.00) | 1.01 | 0 |
| IMCI pneumonia | 0 | 1052 | 100.0% | 0 | 18 | 1.00 (0.81, 1.00) | 0.00 (0.00, 0.00) | 0.02 (0.01, 0.03) |  | 1 |  |
| Danger signs | 3 | 31 | 2.9% | 6 | 6 | 0.33 (0.13, 0.59) | 0.98 (0.96, 0.98) | 0.19 (0.07, 0.37) | 0.99 (0.98, 0.99) | 13.79 (6.45, 29.46) | 0.68 (0.49, 0.95) |
| Danger signs | 2 | 223 | 21.2% | 5 | 11 | 0.61 (0.36, 0.83) | 0.79 (0.77, 0.82) | 0.05 (0.02, 0.09) | 0.99 (0.98, 1.00) | 2.98 (2.02, 4.39) | 0.49 (0.27, 0.87) |
| Danger signs | 1 | 699 | 66.4% | 5 | 16 | 0.89 (0.65, 0.99) | 0.34 (0.31, 0.37) | 0.02 (0.01, 0.04) | 0.99 (0.98, 1.00) | 1.35 (1.14, 1.59) | 0.33 (0.09, 1.21) |
| Danger signs | 0 | 1052 | 100.0% | 2 | 18 | 1.00 (0.81, 1.00) | 0.00 (0.00, 0.00) | 0.02 (0.01, 0.03) |  | 1 |  |

#### Figure S1

Score distributions overall and by outcome. Note different y-axis scales.

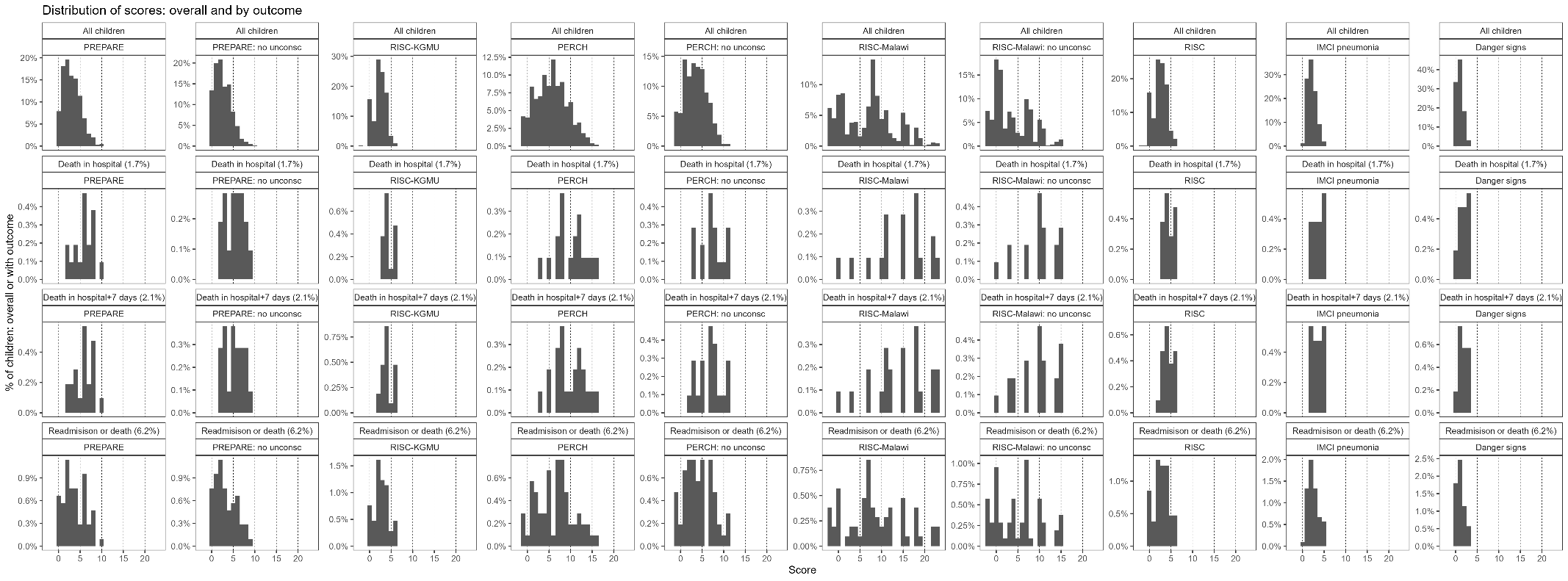

Note: “no unconsc” indicates the PediCAP variable ‘lethargy or unconsciousness’ was considered as ‘lethargy’, rather than as ‘unconsciousness’ as in the primary analysis.

#### Figure S2

Contribution of score components to total score across all children. See table S2 for additional information on score components.

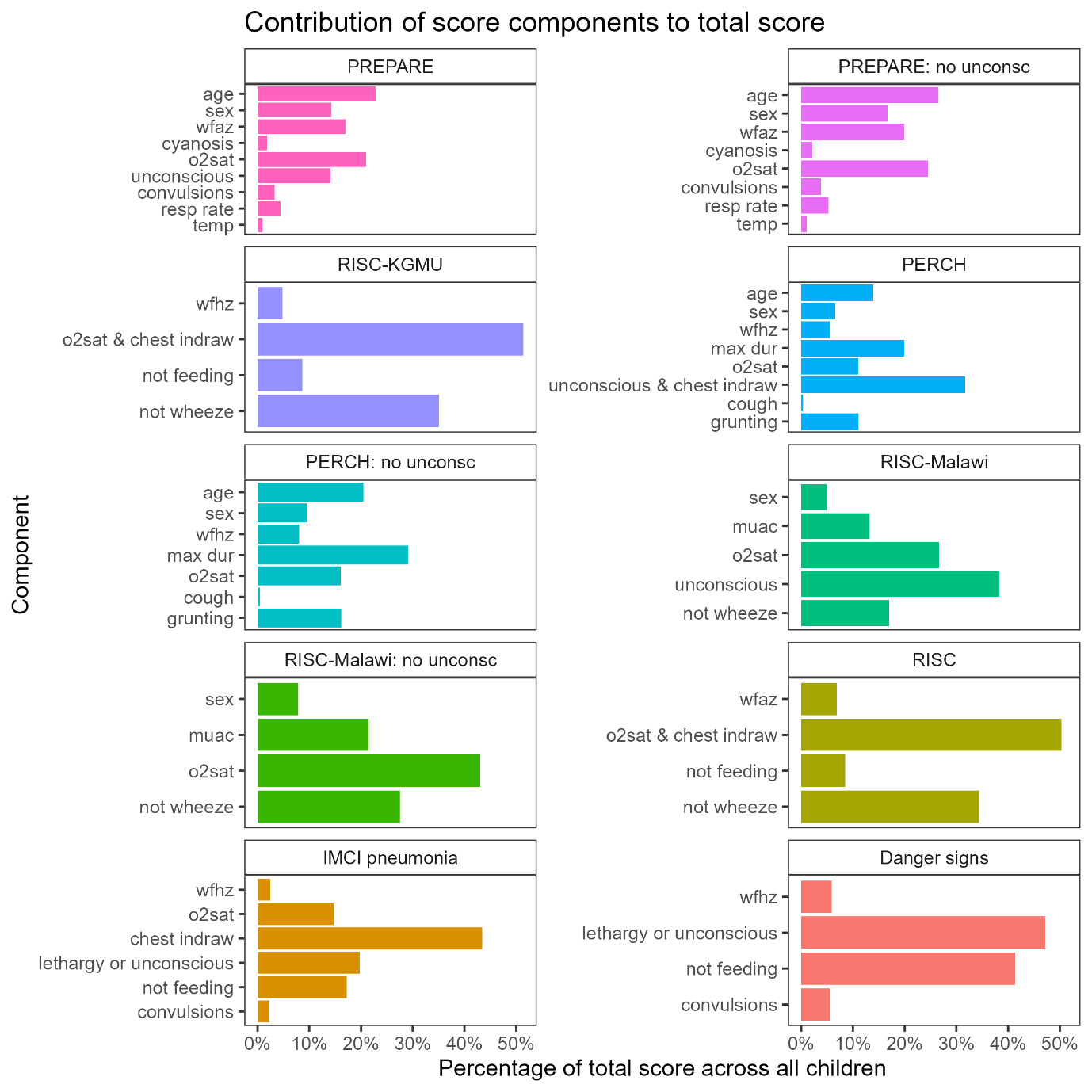

Note: “no unconsc” indicates the PediCAP variable ‘lethargy or unconsciousness’ was considered as ‘lethargy’, rather than as ‘unconsciousness’ as in the primary analysis.

#### Figure S3

Case fatality rates (CFR) and 95% confidence intervals for score components with CFR above the overall CFR for each outcome (red line)

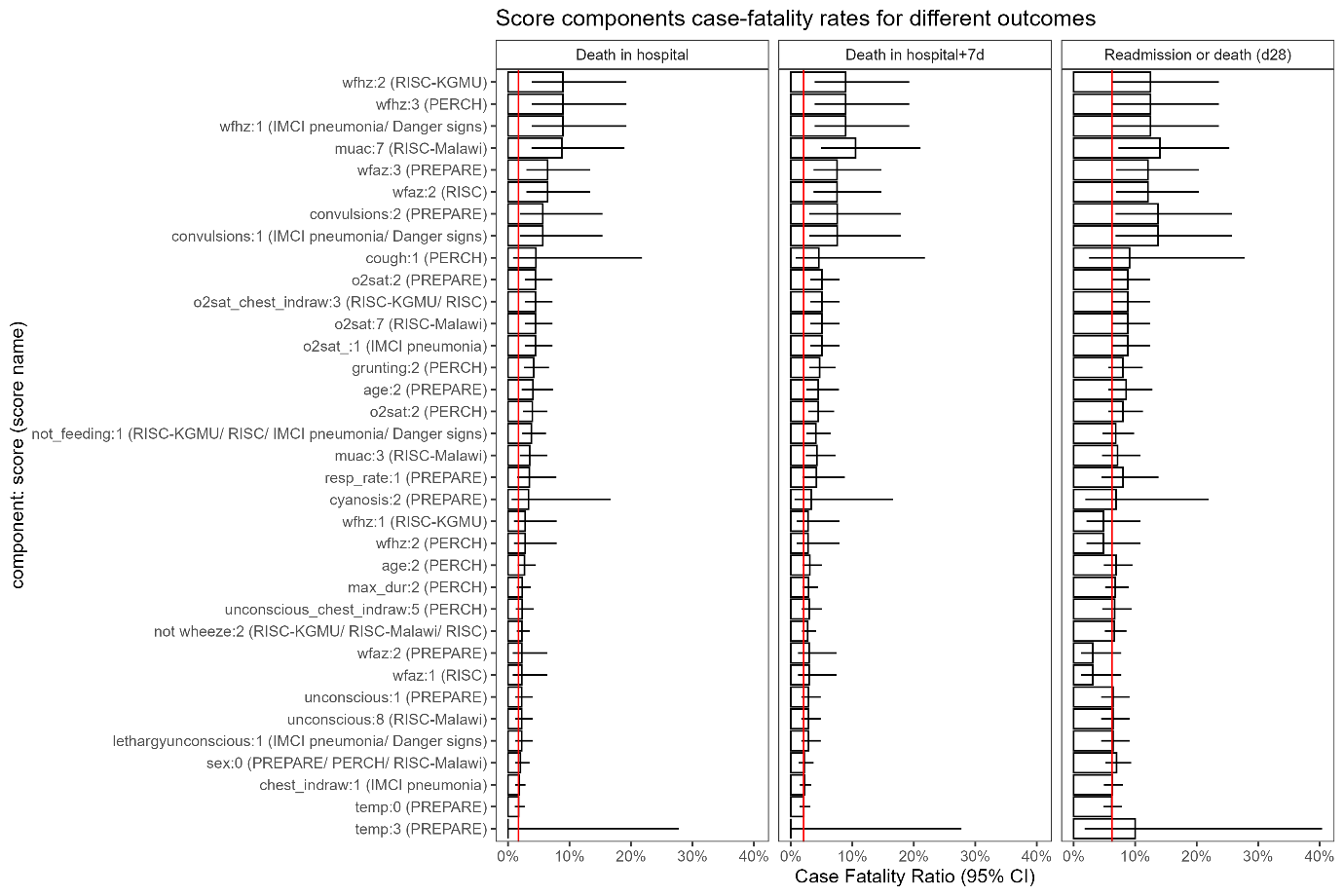

#### Figure S4

ROC-AUC for secondary outcome of death in hospital + 7 days

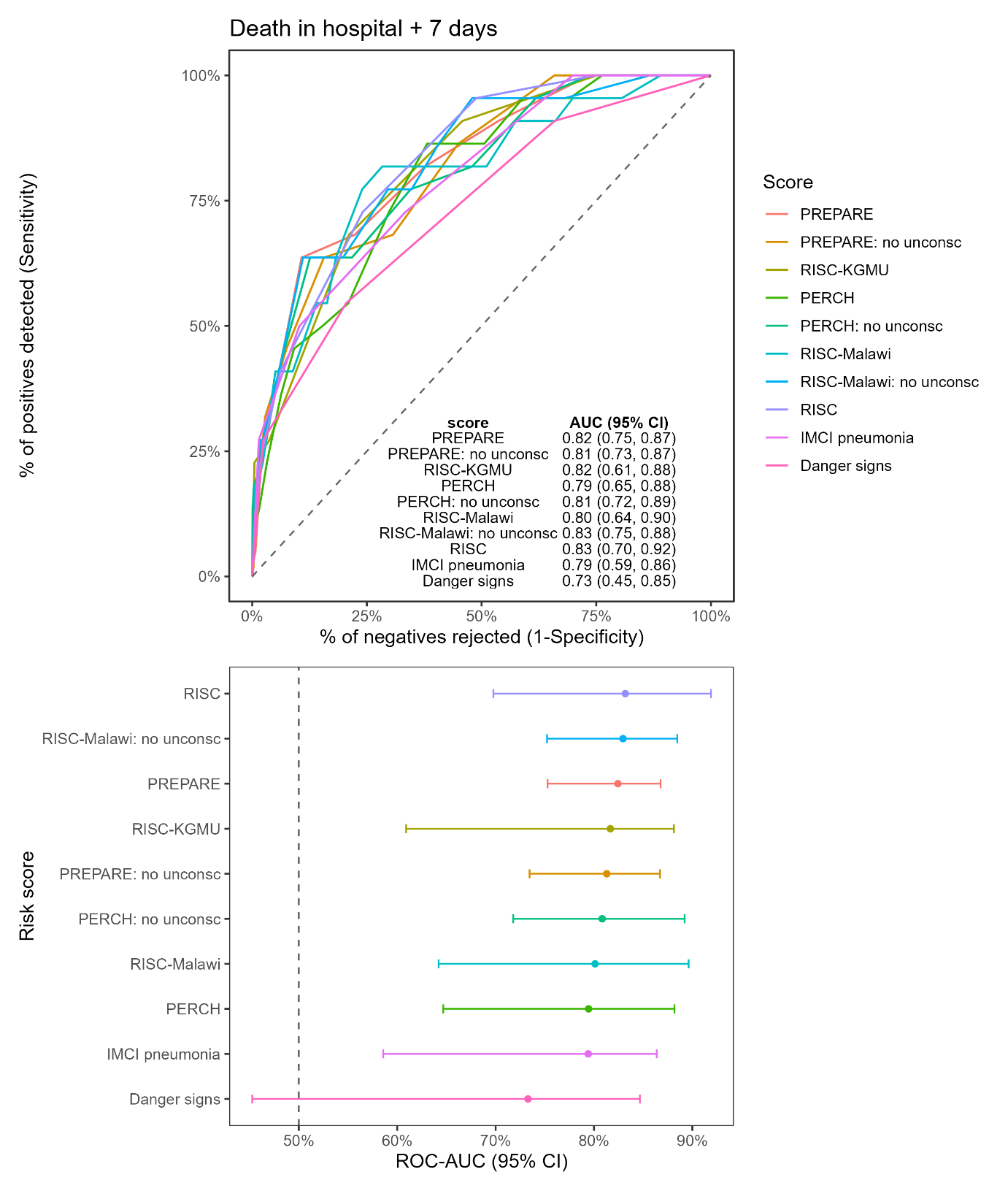

#### Code S1

**R code to calculate performance metrics**

library(tidyverse)

library(pROC) # for calcualting AUC curves

library(gridExtra) # for formatting output

library(PRROC) # for calcualting AUCs

set.seed(1900)

#' reading in files -----------------------------------------

\# read in datasets

scores <- readRDS("03_clean_data/scores_outcomes.RDS")

\# setting vars ------------------------------------------------------------

score_vars <- c("PREPARE", "PREPARE: no unconsc", "RISC-KGMU", "PERCH", "PERCH: no unconsc", "RISC-Malawi", "RISC-Malawi: no unconsc", "RISC", "IMCI pneumonia", "Danger signs")

outcome_vars <- c("deathinhosp", "deathin7days", "readmission_or_death" )

\# calcuating AUC ----------------------------------------------

\# calculate metrics

calc_auc_boot <- function(data,

outcome,

score,

site_var = "Centre",

n_boot = 1000) {

\# -----------------------------

\# point estimates

\# -----------------------------

truth <- data[[outcome]]

preds <- data[[score]]

\# remove missing outcome or score values

keep <- !is.na(truth) & !is.na(preds)

data <- data[keep, ]

truth <- data[[outcome]]

preds <- data[[score]]

auc_val <- tryCatch({

as.numeric(

pROC::auc(

pROC::roc(

truth,

preds,

quiet = TRUE,

direction = "<"

)

)

)

}, error = function(e) NA_real_)

prauc_val <- tryCatch({

PRROC::pr.curve(

scores.class0 = preds[truth == 1],

scores.class1 = preds[truth == 0],

curve = FALSE

)$auc.integral

}, error = function(e) NA_real_)

\# -----------------------------

\# cluster bootstrap by site

\# -----------------------------

sites <- unique(data[[site_var]])

boot_stats <- purrr::map_dfr(

seq_len(n_boot),

function(i) {

if(i %% 100 == 0) {

cat("Bootstrap", i, "of", n_boot, "\n")

flush.console()

}

sampled_sites <- sample(

sites,

size = length(sites),

replace = TRUE

)

boot_dat <- purrr::map2_dfr(

sampled_sites,

seq_along(sampled_sites),

function(s, id) {

data %>%

dplyr::filter(.data[[site_var]] == s) %>%

dplyr::mutate(boot_cluster = id)

}

)

y <- boot_dat[[outcome]]

x <- boot_dat[[score]]

keep <- !is.na(y) & !is.na(x)

y <- y[keep]

x <- x[keep]

auc_b <- tryCatch({

if(length(unique(stats::na.omit(y))) < 2) {

NA_real_

} else {

as.numeric(

pROC::auc(

pROC::roc(

y,

x,

quiet = TRUE,

direction = "<"

)

)

)

}

}, error = function(e) NA_real_)

prauc_b <- tryCatch({

if(length(unique(y)) < 2) {

NA_real_

} else {

PRROC::pr.curve(

scores.class0 = x[y == 1],

scores.class1 = x[y == 0],

curve = FALSE

)$auc.integral

}

}, error = function(e) NA_real_)

tibble::tibble(

auc = auc_b,

prauc = prauc_b

)

}

)

\# remove failed bootstrap iterations

boot_stats <- boot_stats %>%

dplyr::filter(!is.na(auc))

tibble::tibble(

n = nrow(data),

events = sum(data[[outcome]] == 1, na.rm = TRUE),

auc = auc_val,

auc_lo = quantile(

boot_stats$auc,

0.025,

na.rm = TRUE

) |> unname(),

auc_hi = quantile(

boot_stats$auc,

0.975,

na.rm = TRUE

) |> unname(),

prauc = prauc_val,

prauc_lo = quantile(

boot_stats$prauc,

0.025,

na.rm = TRUE

) |> unname(),

prauc_hi = quantile(

boot_stats$prauc,

0.975,

na.rm = TRUE

) |> unname(),

n_boot_valid = nrow(boot_stats)

)

}

\# apply to all outcome-score pairs

results <- tidyr::expand_grid(

outcome = outcome_vars,

score = score_vars

) %>%

mutate(

metrics = purrr::map2(

outcome,

score,

~ calc_auc_boot(

scores,

.x,

.y,

site_var = "Centre",

n_boot = 500

)

)

) %>%

tidyr::unnest(metrics)
